# Adulthood asthma as a consequence of childhood adversity: a systematic review of epigenetically affected genes

**DOI:** 10.1101/2021.07.01.21259888

**Authors:** Yasemin Saygideğer, Hakan Özkan, Oya Baydar, Özge Yılmaz

## Abstract

There is an accumulating data that shows relation between childhood adversity and vulnerability to chronic diseases as well as epigenetic influences that in turn give rise to these diseases. Asthma is one of the chronic diseases that is influenced from genetic regulation of the inflammatory biomolecules and therefore the hypothesis in this research was childhood adversity might have caused epigenetic differentiation in the asthma related genes in the population who had childhood trauma. To test this hypothesis, the literature was systematically reviewed to extract epigenetically modified gene data of the adults who had childhood adversity, and affected genes were further evaluated for their association with asthma. PRISMA guidelines were adopted and PubMed and Google Scholar were included in the searched databases, to evaluate epigenetic modifications in asthma related genes of physically, emotionally or sexually abused children. After retrieving a total of 12,085 articles, 36 of them were included in the study. Several genes and pathways that may contribute to pathogenesis of asthma development, increased inflammation or response to asthma treatment were found epigenetically affected by childhood traumas. Childhood adversity, causing epigenetic changes in DNA, may lead to asthma development or influence the course of the disease and therefore should be taken into account for the prolonged health consequences.

## INTRODUCTION AND AIM

Chronic respiratory diseases, particularly asthma, is known to be regulated by cellular and immunologic responses to environmental and biological factors to varying degrees in different individuals. Prenatal in-utero stress, microbiota at birth or various postnatal factors such as nutrition, infections and second-hand smoke contribute to the development of these responses^1,2^. Cumulative data also indicate a relation between early life adversities and chronic airway diseases. A summary of 8 population-based research articles that show association between respiratory diseases and childhood adversities are listed in **Table I** ^3-10^. Six of these studies are cross-sectional and two of them are case control. These studies not only focus on asthma, but also other chronic disorders including COPD, cancer and immune system disorders. They have evaluated different types of adversities ranging from sexual and physical abuse to gun violence and emotional abuse in children, and the outcome revealed that there was a strong correlation between childhood adversities and presence of adulthood chronic respiratory diseases.

**Table I.** Studies introduce relation with child abuse and respiratory diseases.

| Author,<br>Year | Method | Sample Size | Trauma Type | Significant outcome<br>for respiratory<br>system |
| --- | --- | --- | --- | --- |
| Shields, 2016 [3] | Cross-sectional | 15902 | Childhood physical abuse, sexual abuse, | COPD |
| Bahn, 2014 [4] | Cross- sectional | 84786 | Child sexual abuse<br>And physical abuse | Asthma |
| Ayaydin,2016 [5] | Case-control | 43 | Child sexual abuse | Immune system<br>disorders |
| Remigio-Baker<br>2015[6] | Cross-sectional | 3363 | Physical abuse<br>Verbal abuse<br>Sexual abuse | COPD<br>Asthma |
| Clemens, 2018 [7] | Cross-sectional | 2510 | Child maltreatment<br>(sexual, emotional, physical bause,<br>neglect) | Obesity<br>COPD |
| Salazar, 2016 [8] | Case-control | 747 | Gun violence | Asthma |
| Bonfim, 2015 [9] | Cross-sectional | 1370 | Intrafamilial violence | Asthma |
| Bellis, 2014[10] | Cross-sectional | 3885 | Child abuse | Cancer<br>Resp. disease |

Child adversities are known as risk factors for psycho-social or mental disorders in different levels and found associated with addiction to smoke, alcohol or drugs^11^. In this context, search for the epigenetic changes in acute and chronic stress including childhood adversities revealed that the methylation status of various genes changes due to these childhood conditions^12^. The proposed pathways regarding epigenetic changes suggest that; stress activates hypothalamic pituitary adrenal axis (HPA axis) and this induce cortisol increase leading to direct or indirect regulation of immune system.

The hypothesis of this research was, epigenetic changes might lead to asthma in abused children. Therefore, we systematically screened the studies that listed significantly affected genes from epigenetic changes associated with child abuse and then further evaluated for the role of these genes in asthma pathogenesis.

## METHODS

We followed PRISMA Statement^13^ for review of the literature for childhood adversity and asthma in population-based researches. We used keywords (“child abuse” OR “child adversity” OR “child sexual abuse” OR “child physical abuse” OR “child neglect” OR “child trauma”) AND (“genetics” OR “epigenetics” OR (DNA Methylation)) in NCBI-PubMed and Google Scholar to bring together the informative data that shows relation between childhood adversity and epigenetic changes. Only original research articles were included; review articles, articles involving adversities that are occurred after age of 18 and articles that did not have the results of epigenetic analysis as well as the ones involving only economical adversities and pregnancy adversities were excluded. Initially, 12,085 articles were retrieved, and majority of those articles were excluded after reviewing the abstracts and titles. Among the 36 articles that were identified after this first review, 15 had whole genome methylation arrays and 21 had methylation data for single genes or pathways, given in the adopted PRISMA flowchart (Figure-1). Finally, we extracted significantly methylated genes from each study and evaluated the functions of these genes on G*ene Cards* and NCBI Gene web sites in order to identify the relations with asthma. We also searched PubMed to assess in-vivo, in-vitro and clinical studies in relation with these differentially methylated genes and asthma. We then run a gene ontology analysis using an online tool^14^ and visualized the pathways related with the evaluated genes.

**Fig. 1.**
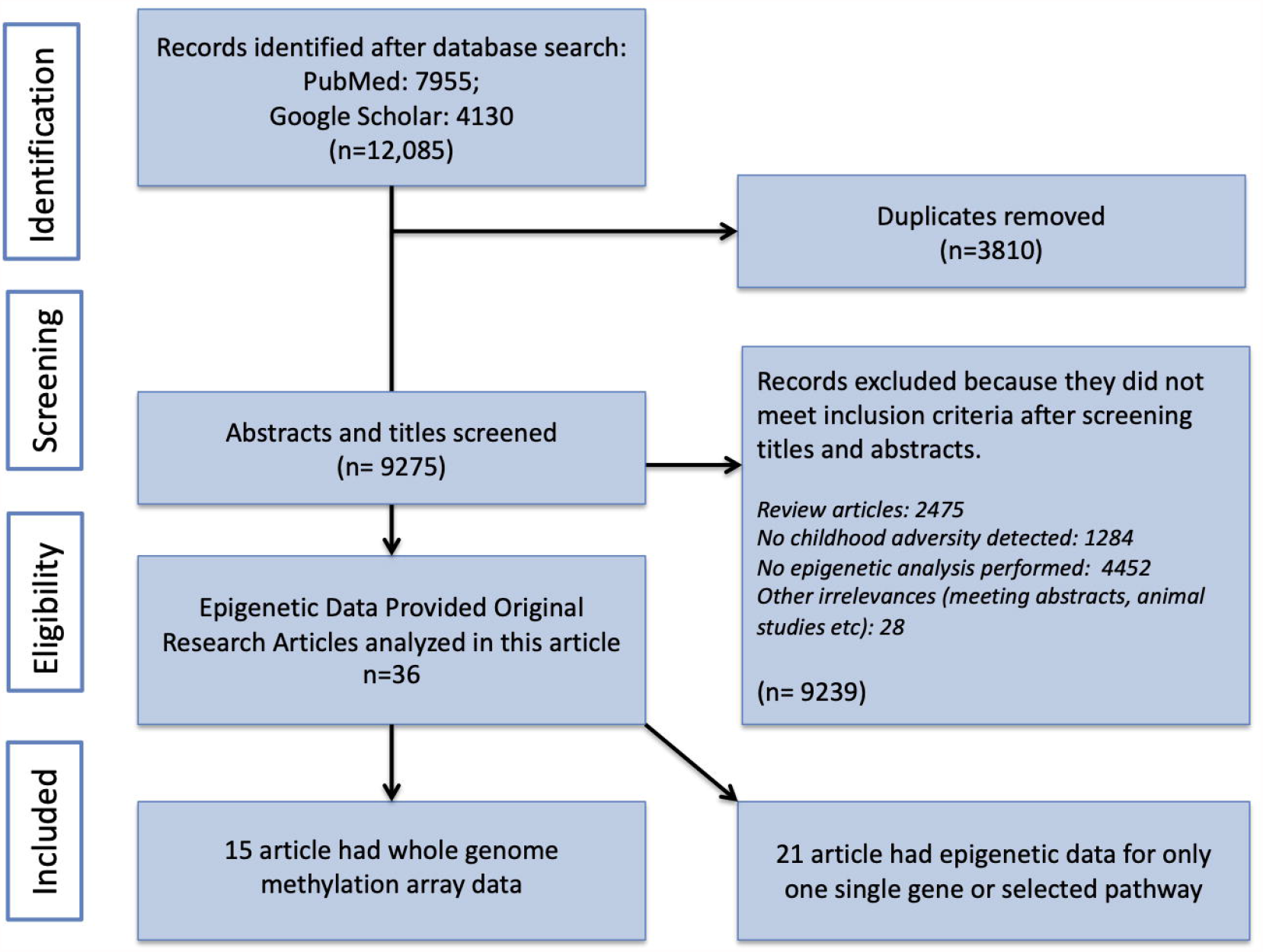
Adopted PRISMA Flowchart as of date 18 Decemeber 2019.

## RESULTS

After reviewing 36 full text articles, differentially methylated genes were recorded and analyzed for their role in asthma. The study sample sizes, methods, studied tissues, countries, and significantly differently methylated genes and their possible contribution to asthma is listed in Table-II for 15 whole genome analysis or microarray studies. Twenty-two single gene or pathway studies are listed in supplementary table. All studies were performed in developed countries including USA, Canada, UK, and Netherlands etc. and they have extracted DNAs were from whole blood, saliva, T cells and monocytes. There was at least one affected gene related to asthma in each study while some of the studies showed multiple pathways involved in asthma pathogenesis affected (Table-II). Most studies had data for post-traumatic stress disorder genes and focused on hypothalamus -pituitary – adrenal (HPA) axis and immune system related genes, therefore, the genes that regulate HPA axis, *NR3C1* and *FKBP5*, were the most well-studied genes. *NR3C1* and *FKBP5 are also involved* in asthma pathogenesis by decreased suppression of inflammation and decrease response to corticosteroids respectively. Other significantly deregulated genes that were affected by childhood adversities were interleukins, ADAM family, WNT, STAT and MAPK pathway and inflammatory related genes (Table-II). Only one recent study had found no significant relation between childhood adversity and epigenetic regulation after applying multiple statistical corrections (Table-II).

**TABLE-II.**
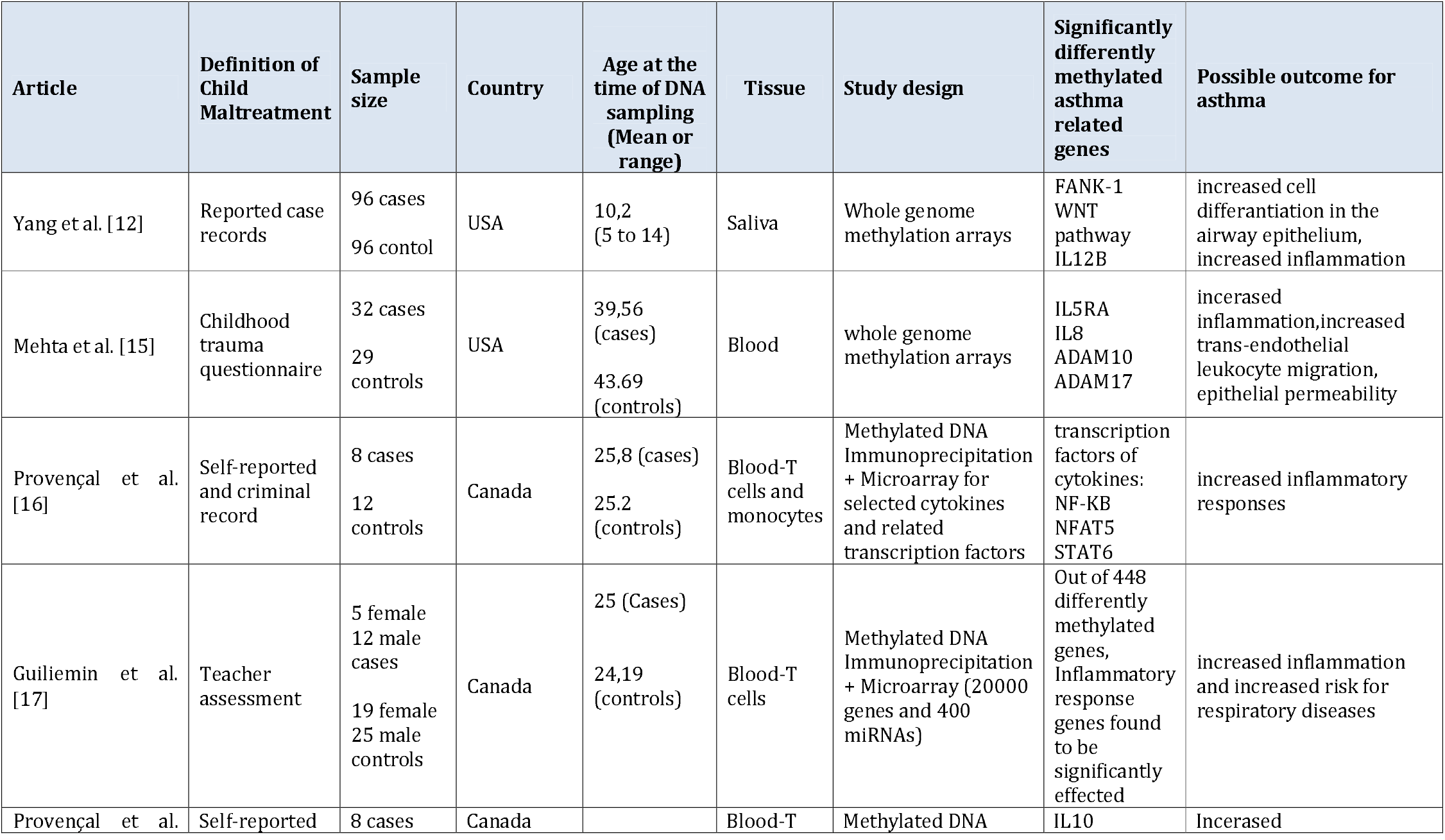
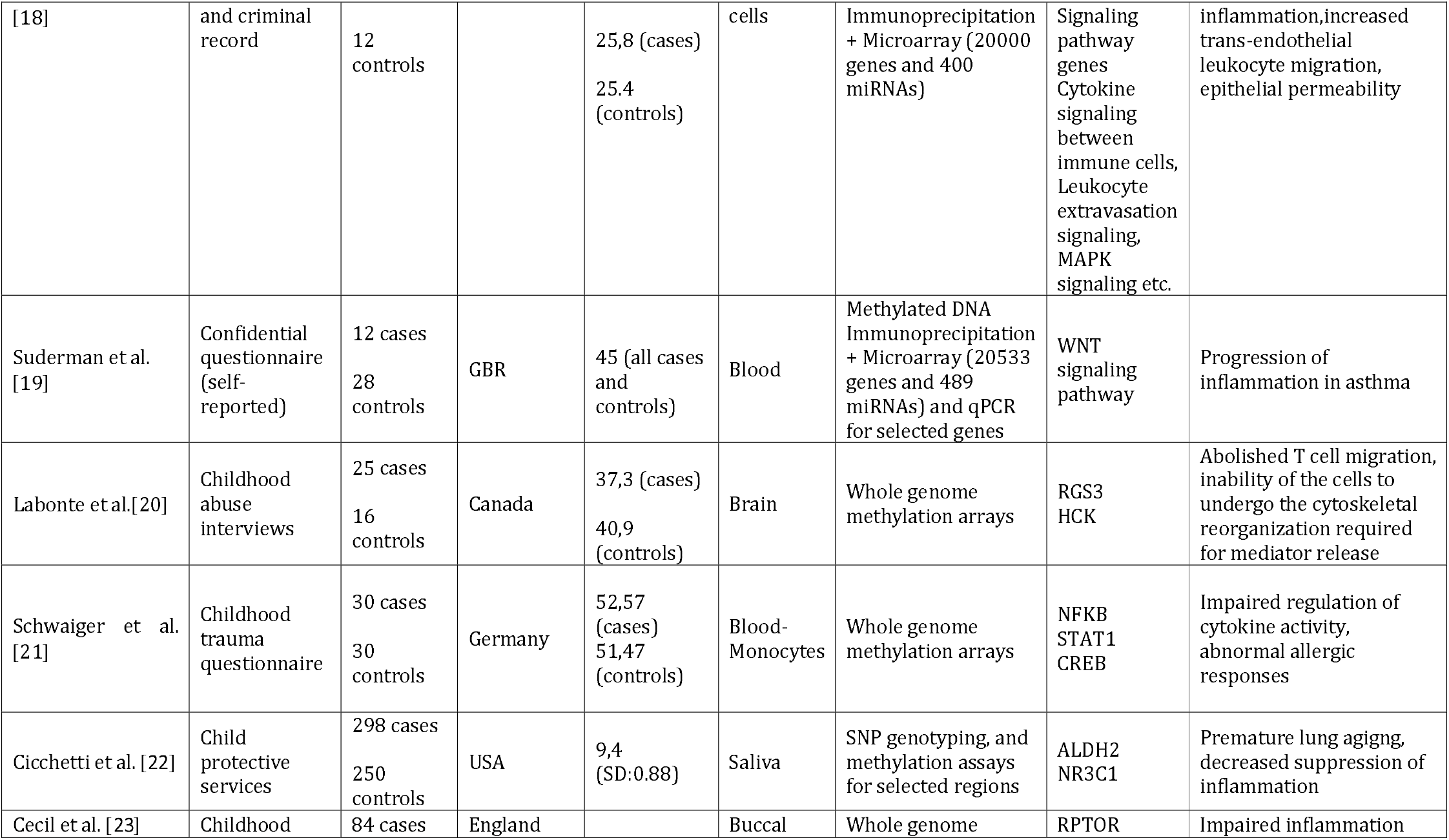
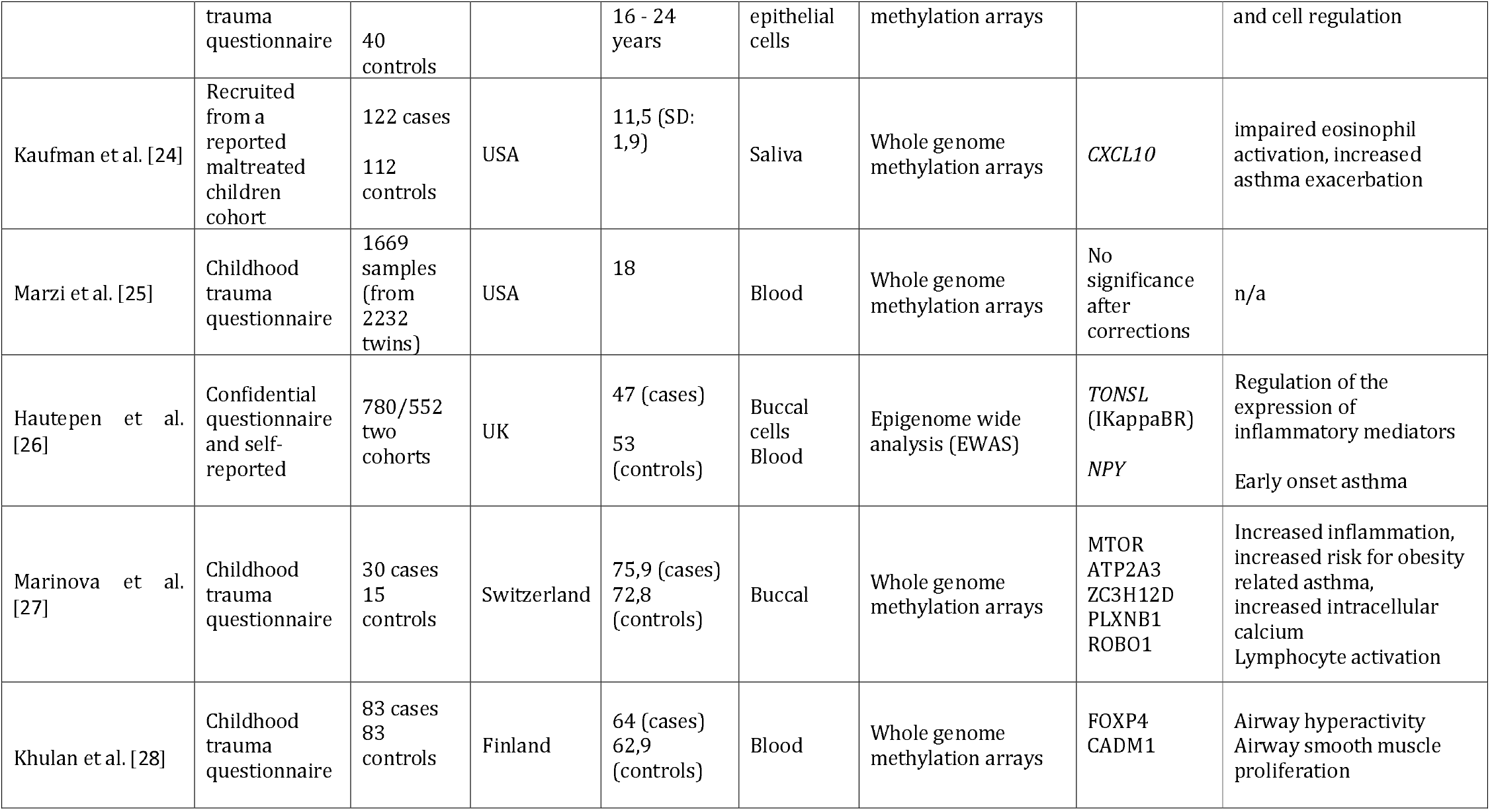
Properties of 15 studies of childhood adversity and possible effects of differentially methylated genes in asthma.

### *NR3C1* and Asthma

The *NR3C1* gene encodes glucocorticoid receptor (GR), which has DNA binding, nuclear localization, ligand binding and two activation function domains, and has the ability to inhibit the expression of asthma related cytokines such as IL-5^29^. IL-5 induces the activity of eosinophils in asthma, after being released from the active macrophages and T lymphocytes. IL-5 induced leukotrienes, therefore, causes asthmatic reactions and symptoms. In a recent bioinformatics study that evaluated ignored genes that might have a function in allergic asthma, *NR3C1* was found to be downregulated^30^. It has also been shown that polymorphisms and mutations in specific sites of this gene has correlation with increase in TGF-β expression in asthma patients^31^, which is a well-known mediator that leads smooth muscle proliferation and airway remodeling. Regarding the epigenetic regulation of *NR3C1*, maternal stress during pregnancy, caused increased methylation in the umbilical cord blood mononuclear cells that cause decreased GR expression in the child, which may affect cytokine production and treatment response in the childhood asthma^32^.

### *FKBP5* and Asthma

FKBP5 or FK506-binding protein 51, is a member of immunophilin protein family that has peptidyl-prolyl isomerases to catalyze isomeric shape of proline amino acid and thus, act as co-chaperon to assist protein folding in various proteins. Increased levels of protein is found in central and peripheric airway brushings of severe asthma patients comparing to healthy subjects^33^. It also serves as a receptor for steroids and immune suppressive drugs and therefore, mostly studied for the effects of treatment response in asthma. FKBP5 express an inhibitory effect on GR function by binding to glucocorticoid-GR complex and delays nuclear translocation of the signal^34^. There are also other rules of this protein in different pathways related with asthma pathogenesis. Increased levels of FKBP5 induce T-cell proliferation and function by binding to calcineurin and activating NFAT (Nuclear factor of activated T-cells) pathway^34^. Besides, stress induced decrease in *FKBP5* methylation, upregulated the protein levels in blood and immune cells and this increase in FKBP5 promotes NF-kB (nuclear factor kappa-light chain enhancer of activated B cells) mediated inflammation^35^. Thus, reduced methylation of FKBP5 in the abused children, might play role in the asthma development.

### *FANK1* and Asthma

The relationship of FANK1 (Fibronectin type 3 and ankyrin repeat domains protein 1) with asthma is not well-studied There are studies that suggest the role of FANK1 in inhibition or activation of apoptosis. It is known to be expressed mostly by testis cells, but recently shown to be present also in T cells, alveolar and epithelial cells of bronchi^36^. In a computational analysis of asthma related genes, *FANK1* has come forward as one of the 10 genes found to be associated with asthma^37^. However, the functional relation currently remains unknown.

### *CHRNA5* And Asthma

*CHRNA5* (Neuronal acetylcholine receptor subunit alpha-5), is a cholinergic nicotinic receptor gene that acts in opening ion channels on the plasma membrane after acetylcholine binding. Expression of the gene is mostly found in airway myocytes and epithelial cells throughout the lungs^38^. In a meta-analysis that evaluate chromosome 15q25 region and airflow obstruction, Asp389Asn missense single nucleotide polymorphism in *CHRNA5* was associated with airflow obstruction in never smokers^38^. Silencing this gene in three-dimensional cell culture model, increased thickness of the epithelium, suggesting the lead to airway remodeling^39^. Studies show increased dependency to cigarette smoking in men^40-41^ but there is no data that shows relation with epigenetic regulation of *CHRNA5* and asthma but since genetically modification of the gene contributes airflow obstruction, and in vitro knock-down of the gene causes increased thickness in the airway models, it is possible that the increased methylation might contribute to asthma phenotype in the abused child.

### *ADAMs 10 and 17* and Asthma

ADAM (A disintegrin and metalloproteinase domain containing protein) family proteins act as sheddase, with its adhesion and protease function, and shed various membrane proteins at the outer cell membrane that cause the maturation of those proteins and/or releasing them from the membrane. A single ADAM protein can cleave more than one protein as well as different ADAM members may cleave the same substrate. ADAM10 and ADAM17 have the similar active sites that both of them can cleave membrane bound TNF-alpha to its mature dissolvable state. ADAM10 has ability to shed ephrin/eph complexes between the cells, helps releasing soluble forms of IL6 and IL11 by mediating cleavage of IL6R and IL11RA, and also other surface proteins including heparin binding epidermal growth factor. ADAM10 also serves as a receptor for S. aureus, increasing the virulence of the bacteria. ADAM17 also cleaves IL6R, IL1RII, TGF-alpha, growth hormone receptor^42^. Obviously, there are other many proteins and pathways effected by these two ADAMs that are not listed here. Thus, ADAM 10 and 17 playing role in endothelial permeability, smooth muscle transactivation, leukocyte recruitment, preventing resolution of inflammation and inducing inflammation, are important proteins in asthma pathogenesis and might be target for the treatment of the disease. Loss of function mutations of *ADAM10* and *ADAM17* are extremely rare and mice with double knockout of these genes do not survive^42^. Overall, epigenetic regulation of these genes might affect asthma development in children.

### *SLC6A4 (SERT)* and Asthma

*SLC6A4* gene, encodes sodium dependent serotonin transporter and solute carrier family 6-member 4 (also SERT or 5HT transporter) protein that involves in recycling of serotonin (5-Hydroxy tryptamine, 5HT) molecules by transporting them from the synaptic area to the pre-synaptic membrane. 5HT, besides its functions in the central nervous system, has ability to induce cytokine and chemokine synthesis, cell proliferation and tissue regeneration^43^, therefore, it is well studied in the pathogenesis of asthma. Increased levels of serotonin is found in symptomatic asthmatic patients and negatively correlated with FEV1 levels^43^. 5HT, stimulates PGE2 production by alveolar macrophages, activates eosinophils and suppress IL-12 expression^43^. 5HT also act as a platelet-stored vasoconstrictor. Termination of the serotonin signal depends on the levels of SLC6A4 and activation of the receptor helps relief of asthma symptoms to release^44^. There are studies that evaluated the relationship between *SLC6A4* gene polymorphisms and asthma but have not found any correlations up to now with the studied ones^45^. *SLC6A4* knockout mice show anxiety-like behavior and changes in the methylation of the gene may contribute the levels of 5HT and therefore influence asthma genotype in the maltreated and affected children.

### *OXTR* and Asthma

*OXTR* gene codes for oxytocin receptor, which is a G protein coupled receptor that needs to activate phosphatidylinositol-calcium second messenger molecules. Studies have shown that OXTR is expressed both in human and mice airway smooth muscle cells and its expression is affected by cytokines such as IL-13 and TNF-alpha^46^. Oxytocine levels in BAL fluids were also found increased in asthmatic patients comparing to healthy control subjects^46^ suggesting that *OXTR* and its epigenetic regulation might have a role in the pathogenesis of asthma.

### *RPTOR, MTOR* and Asthma

RPTOR or raptor (Regulatory-associated protein of mTOR), regulates mammalian target of rapamycin complex 1 (mTORC1) activity which involves in cell growth, survival and autophagy. It also has role in maintaining the size of the cell. It is mostly expressed in muscle cells including lungs. In a study that evaluate airway smooth muscle tissue with RNA sequencing to compare asthmatic and normal people, RPTOR was found one of the four genes which were associated with airway hyper-responsiveness^47^. mTOR (mechanistic target of rapamycin kinase), was also found to involve in airway remodeling models in mouse models as well as regulation of Th17/Treg and Th1/Th2 ratio^48^. Therefore, epigenetic regulation of mTOR and related pathways significantly influence asthma development.

### *RGS3* and *HCK* and Asthma

RGS3 involves in inflammation and T cell migration^49^ and HCK takes plays in MAPK signaling pathway which in turn related with airway smooth muscle proliferation^50^. Labonte et al found these two genes were significantly hypermethylated in the brain tissue of the traumatic children, but currently there is no data that shows the epigenetic change in the blood or other tissues in these two genes.

### *CXCL10* and Asthma

C-X-C motif chemokine ligand 10 (IFN-gamma-induced protein 10 (IP-10) or small inducible cytokine B10) is a small protein coded by *CXCL10* gene. It is expressed in monocytes, endothelial cells and fibroblasts and has a variety of role in the inflammation. Therefore, the effects on the asthma pathogenesis is well-studied. mRNA levels of CXCL10 were found increased in bronchoalveolar fluid of asthmatic patients and data suggest that it has role in corticosteroid therapy induced persistent type I inflammation^51^. Kaufman J et al, evaluated epigenetic influences on the obesity in adverse childhood experienced children and found decreased methylation of the promoter region of *CXCL10*. Therefore, inducing obesity or involving inflammation, epigenetic regulation of the gene may contribute to asthma development in the abused children.

### *STAT* and Asthma

STAT family of proteins (signal transducer and activator of transcription) act as transcription factors after extracellular cytokine or growth factor binding in the cell and lead to the expression of proliferation, differentiation and apoptosis related genes. *STAT1* was reported one of the asthma-ignorome gene together with *NR3C1*^*30*^ and variants of STAT6 found related with risk for asthma in a meta-analysis study^52^. Epigenetic regulation of Jak-STAT pathway in childhood asthma has been recently reported^53^.

### *CREB* and Asthma

CREB (cAMP response element-binding protein) is another transcription factor that regulate expression or suppression of multiple genes after getting activated by upstream signaling pathways those use cAMP as secondary messenger. Therefore, it involves multiple pathways in Asthma signaling including therapy response^54^.

### *NFKB, IKappaBR* and Asthma

NF-KB (nuclear factor kappa-light chain enhancer of activated B cells), consists of protein to regulate transcription, cytokine production and survival when the cell is under stress. Together with MAPK pathway, epigenetic regulation of NF-KB signaling was found significantly affected in adult-onset asthma^55^. Researchers have been interested in targeting NF-KB for asthma treatment for the last years. *IKappaBR (IKBR)* or *TONSL* gene codes for Tonsoku like DNA Repair protein that negatively regulate NFKB mediated transcription. Therefore, increased methylation of IKBR and demethylation of NFKB would worsen asthma phenotype in the maltreated children.

### *ALDH2* and Asthma

ALDH2 (acetaldehyde dehydrogenase 2) is a member of ALDH family of proteins that acts as an antioxidant and its increased expression inhibits the harmful effects of oxidative stress. Polymorphisms of the gene may cause premature lung aging^56^ and ALDH2-deficient mice had alcohol induced asthma in an animal model study^57^. These and several other studies indicate that increased methylation of *ALDH2* may have role in the asthma pathogenesis.

### MAPK pathhway, *NPY - FOXP4* and Asthma

Mitogene activated protein kinase (MAPK) pathway is a series of signaling cascade pathways that induce distinct pathways in the cell regarding differentiation, proliferation and cell survival. Its role in asthma pathogenesis has been shown and reviewed in the literature^58^.

NPY (Neuropeptid Y), involves in multiple signaling pathways including MAPK pathway and it also regulates intracellular calcium levels. The expression levels of NPY on antigen presenting cells (APCs) in allergic asthma has been shown and its role on cytokine expression is not yet clear^59^. In a study, the loss of FOXP4 (Forkhead box protein 4) together with FOXP1, which are transcription factors located in airway cells, led to increased NYP expression resulting in airway hyperreactivity through activation of smooth muscle myosin light-chain phosphorylation^60^.

### *ATP2A3* and Asthma

ATP2A3 (Sarcoplasmic / Endoplasmic Reticulum Calcium ATPase 3), involving intracellular calcium levels, influence a variety of signaling pathways. the expression was shown to vary among different childhood asthma phenotypes^61^.

### Other relevant genes and gene ontology and pathway analysis

The rest of the genes listed in Table-II are usually found in the literature That assessed obesity asthma relationship or that took place in the gene expression pathways in immune system. After reviewing these genes in the literature, we further ran a gene ontology (GO) analysis using two different online tools^62-14^ and visualized the pathways related with the evaluated genes (Figure-2 & Supplementary material-1).

**Fig. 2.**
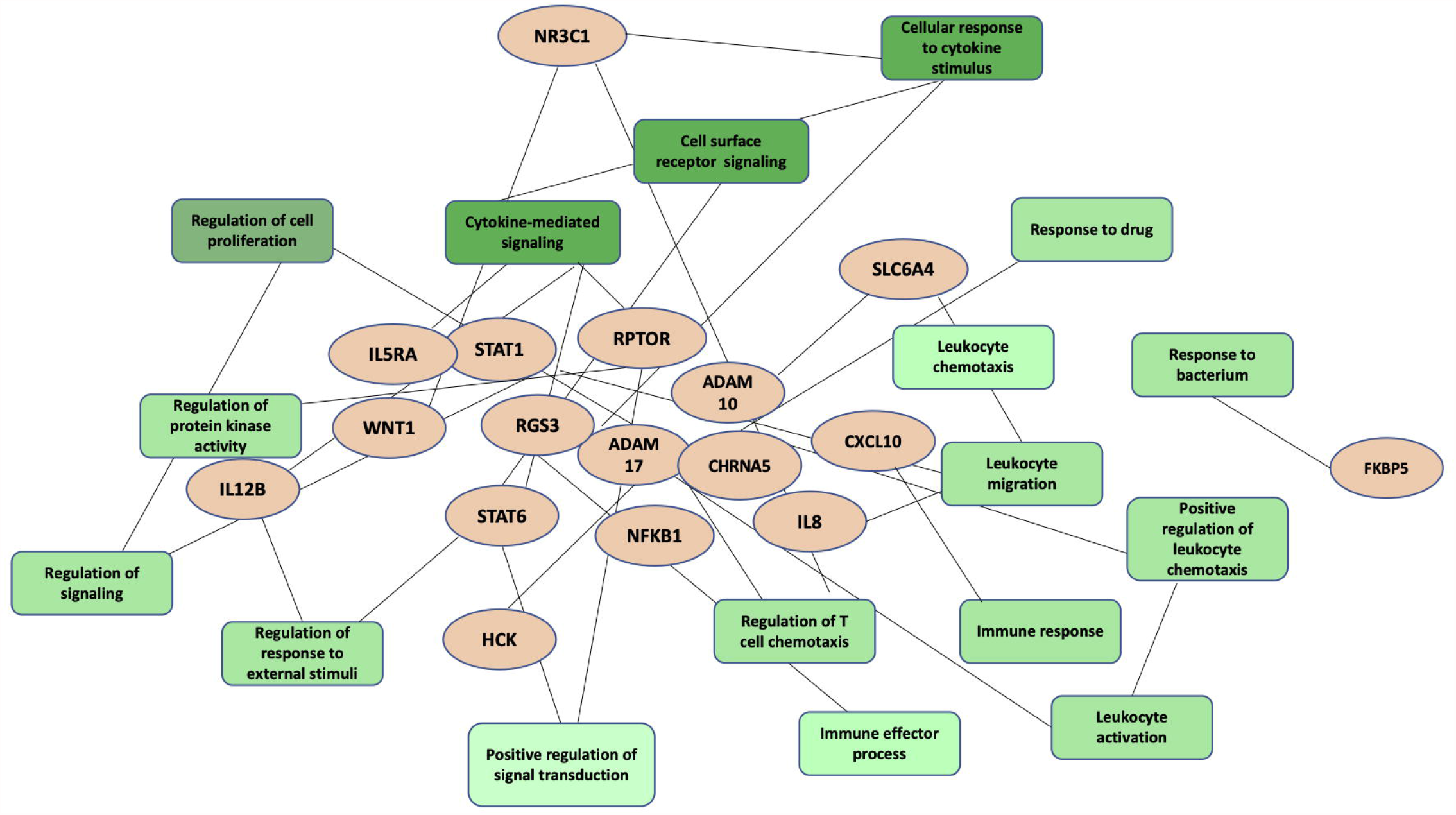
Gene enrichment and pathway analysis of the affected genes. Differentially methylated genes in the adults who had childhood adversity, that have relationship with asthma are shown in orange color and significantly related pathways are in green. Dark green shows strongest relation. The full list of the pathways and corresponding full image are given in supplementary material.

## DISCUSSION & CONCLUSION

In this research, we demonstrated that childhood adversity, including sexual and physical abuse and neglect, effects the epigenetic regulation of asthma related genes. These epigenetic changes focused on altered methylation of DNA, which has become convenient to study comparing to chromatin purification^63^. The increased or decreased methylation of the genes found in the adults, who experienced adverse childhood events, were included well studied asthma related genes such as *ADAM10* &17 and *CXCL10* as well as the genes that seemed less common in asthma pathogenesis regarded studies such as *ALDH2, FOXP4*, and *FANK1*. Up to date scientific investigations of asthma pathogenesis indicate a multi-complex pathophysiologic process that might be responsible of the diverse phenotypes among asthma patients and recent studies indicate a contribution of lifelong epigenetic influences to this pathophysiology^64^. The complexity increases while the type of the adversity and its relationship with asthma may differ from population to population, related to cultures and traditions, and therefore, limitation of this systematic review is that the origin of most studies included the countries with similar socioeconomic background decreasing the value of generalizability. This study also did not include in-utero period of the children, which may also contribute to epigenetic alterations. Although the genes listed in this study have been associated with asthma and a few of them are well-studied, the underlying pathway for these associations may need deeper research for future discoveries of personalized medicine and to understand the pathogenesis of the relationship of these epigenetic influences with asthma.

## Data Availability

Data is available upon request

## Author Contributions

YS designed the study analyzed the results and wrote the manuscript; HO and OB involved in literature search and data collection; OY critically reviewed the manuscript.

